# CESCProg: A COMPACT PROGNOSTIC MODEL AND NOMOGRAM FOR CERVICAL CANCER BASED ON miRNA BIOMARKERS

**DOI:** 10.1101/2023.03.22.23287522

**Authors:** Sangeetha Muthamilselvan, Ashok Palaniappan

## Abstract

Cervical squamous cell carcinoma, more commonly cervical cancer, is the fourth common cancer among women worldwide with substantial burden of disease, and less-invasive, reliable and effective methods for its prognosis are necessary today. Micro-RNAs are increasingly recognized as viable alternative biomarkers for direct diagnosis and prognosis of disease conditions, including various cancers. In this work, we addressed the problem of systematically developing an miRNA-based nomogram for the reliable prognosis of cervical cancer. Towards this, we preprocessed public-domain miRNA -omics data from cervical cancer patients, and applied a cascade of filters in the following sequence: (i) differential expression criteria with respect to controls; (ii) significance with univariate survival analysis; (iii) passage through dimensionality reduction algorithms; and (iv) stepwise backward selection with multivariate Cox modeling. This workflow yielded a compact prognostic DEmiR signature of three miRNAs, namely hsa-miR-625-5p, hs-miR-95-3p, and hsa-miR-330-3p, which were used to construct a risk-score model for the classification of cervical cancer patients into high-risk and low-risk groups. The risk-score model was subjected to blind validation on an unseen test dataset, yielding a one-year AUROC of 0.84 and five-year AUROC of 0.71. The model was validated with an out-of-domain, external dataset yielding significantly worse prognosis for high-risk patients. The risk-score was combined with significant features of the clinical profile to establish a validated predictive prognostic nomogram. Both the miRNA-based risk score model and the integrated nomogram are freely available for academic and not-for-profit use at CESCProg, a web-app (https://apalania.shinyapps.io/cescprog).

## INTRODUCTION

Cervical cancer (cervical squamous cell carcinoma; CESC) ranks fourth globally among cancers in women, and second among women of reproductive age. Due to unequal implementation of invasive screening techniques, the morbidity and mortality rate of cervical cancer continues to rise in countries like India, where it accounted for 9.4% of all cancers and 18.3% of new cases in 2020^1^. Multiple etiological factors contribute to its incidence, including persistent infection of human papilloma virus (HPV)^2^, and known lifestyle factors such as excessive smoking and use of contraceptive pills. Cervical cancer tends to be refractory to treatment unless detected early, and its prognosis is vital to quality-of-life expectations. Late diagnoses in the advanced stages of cervical cancer require expensive and complex treatment, with concomitant poor prognoses^3^. Many gaps remain with respect to cervical cancer screening, diagnosis and prognosis^4^, and biomarkers with high specificity and sensitivity are necessary.

MiRNAs exert key control over regulation of gene expression^5^, by inducing specific translational repression via target mRNA 3′ UTR deadenylation and decapping. MiRNAs are known to target ∼60% of the transcriptome, thus modulating biological processes^6^. Their aberrant, differential expression is implicated in various cancers, where they act as either oncogenes (oncomirs) or tumor suppressor genes (mirsupps), regulating tumorigenic process like cell maturation, cell proliferation, migration, invasion, apoptosis, and metastasis^7^. MiRNA biomarkers from the serum or cervical mucus could potentially augment systems for early diagnosis, prediction of disease progression, and outcome improvement, in addition to facilitating prognostic information, with respect to cervical cancer. The US National Cancer Institute launched the Cancer Genome Atlas (TCGA) to characterize different tumor types using -omics platforms, and make raw and processed data available to all researchers^8^. In this study, we used the TCGA CESC miRNA -omics dataset to build a validated prognostic risk model and predictive nomogram based on a minimal miRNA signature and the clinical profile. The developed models have been deployed as a freely-available web-app service for non-commercial use at CESCProg (https://apalania.shinyapps.io/cescprog).

## MATERIALS AND METHODS

The workflow is summarised in Fig. 1, and discussed in detail below.

**Figure 1.**
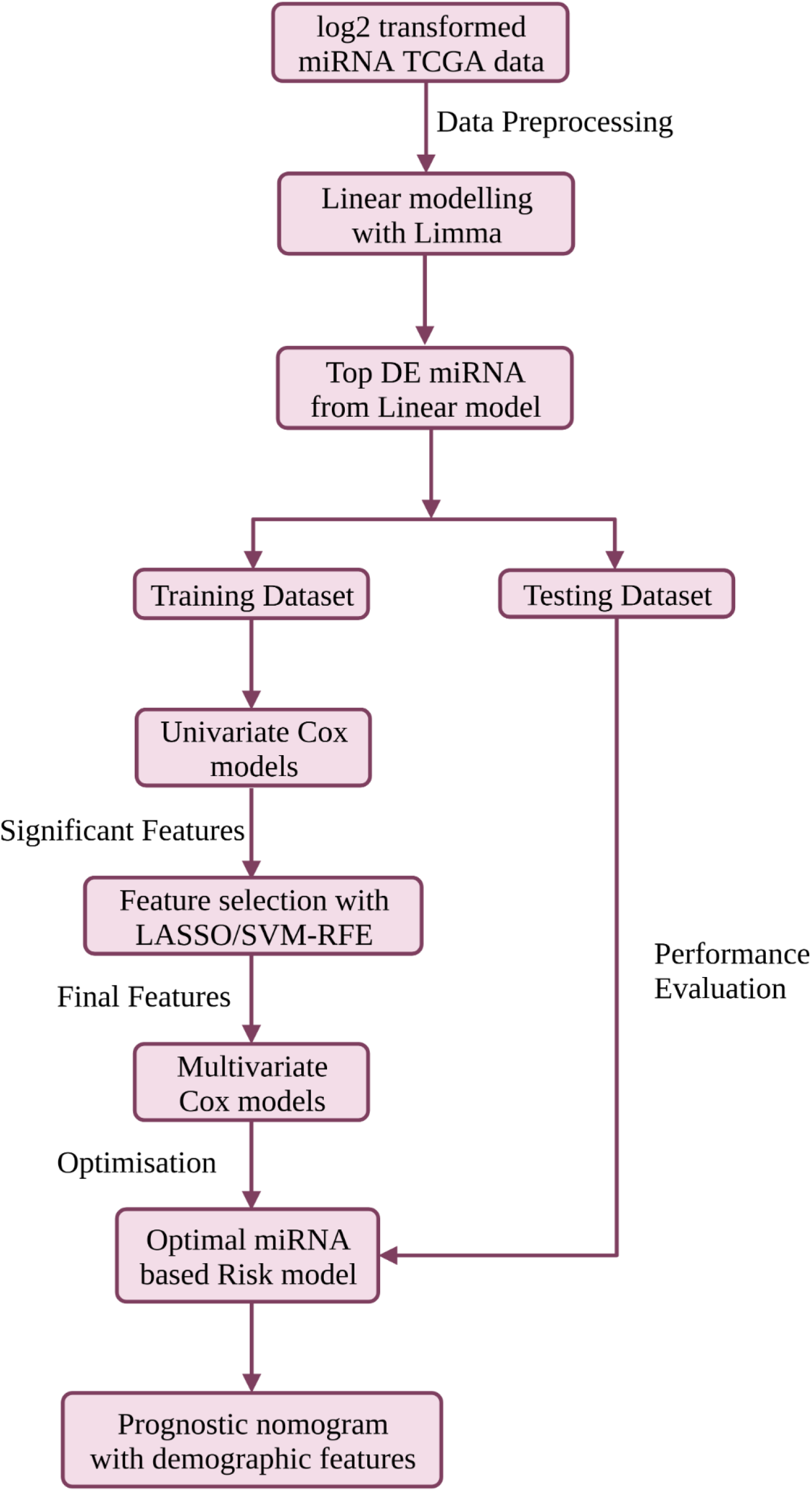
The workflow used in this study for the development of a compact validated risk model for cervical cancer prognosis. The predictive prognostic nomogram was re-built with the full dataset prior to deployment at CESC-PROG (https://apalania.shinyapps.io/cescprog).

### Data preprocessing

Normalized and log_2_-transformed Illumina HiSeq miRSeq data preprocessed with the TCGA miRNA analysis pipeline were obtained from firebrowse.org portal^9^. The patient barcode of each sample was parsed to annotate the samples as ‘normal’ and ‘cancer’. The corresponding clinical metadata was also retrieved from firebrowse.org (CESC.Merge_Clinical.Level_1.2016012800.0.0.tar) and used to annotate the stage information (encoded in ‘patient.stage_event.clinical_stage’ variable) of the tumor samples, and then merged with the expression data. The clinical stage is essentially the surgical stage prior to any treatment received, from the biopsy obtained at the time of surgery. Collapsing possible substages (A, B, C) in each stage yielded the four-class macro-progression of stages (I, II, III, IV). Certain demographic and clinical factors in the metadata including age, HPV status, smoking history, pregnancies, histologic grade, vital status and were retained. Based on the merged dataset, miRNAs with negligible change in expression across samples (expression σ < 1) were removed, as were samples with absent stage information. R (www.r-project.org) was used for dataset preprocessing.

### Linear modelling

The miRNA expression analyses of cancer stages relative to the normal tissue (controls) were performed using the limma package in R^10^. The workflow was essentially adapted from earlier protocols developed in our lab^11^. To recapitulate, a linear model was fit using controls as intercept and sample stages as indicator variables. The fit model was adjusted with empirical Bayes to obtain moderated t-statistics^12^. Multiple hypothesis testing and the false discovery rate were applied using the method of Hochberg and Benjamini to yield adjusted p-values of the F-statistic of the linear fit^13^. Based on the fold change (FC) in the expression of individual miRNAs across conditions, miRNAs with |log2(FC)| > 2.0 and adj. p-value < 0.05 were considered significantly differentially expressed miRNAs (DEmiRs). The preprocessed dataset was then split into train and test datasets in the ratio 0.8:0.2. The test dataset was used for the performance evaluation of the final model, but kept invisible to the model development process.

### Development of compact miRNA signature

Univariate Cox models^14^ were used to screen the DEmiRs by significance, and only DEmiRs with p-value < 0.05 were filtered for further analysis. Two robust feature selection methods, namely Least absolute shrinkage and selector operation (LASSO) Cox regression^15^ and Support vector machine - recursive feature elimination (SVM-RFE)^16^, were used in combination to reduce the dimensionality of the prognostic DEmiRs. LASSO, a form of ‘penalized’ regression with L1 penalty, was implemented using R-glmnet^17^, whereas SVM-RFE, which computes ranking weights for all features and then iteratively performs backward selection, was implemented using R-e1071^18^. A union of the features selected from these two implementations was taken forward and used in a stepwise multivariate Cox logistic regression^19^ for establishing the prognostic DEmiR signature of cervical cancer.

### Prognostic risk model

A risk model was formulated based on the identified prognostic DEmiR signature and used to evaluate the survival risk of each patient. It is given by the exponent in the multivariate Cox model:

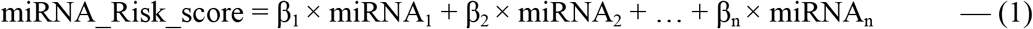

where n is the size of the prognostic DEmiR signature, miRNA_i_ denotes the expression level of the i^th^ miRNA, and β_i_ denotes the effect-size (or weight) of the i^th^ miRNA. Applying the optimal cut-point (i.e, median) given by maxstat (maximally selected rank) statistic from the R-survminer^20^ to the risk score distribution, we categorized (binarized) patients with CESC into high-risk and low-risk groups. Kaplan-Meier curves and AUROC were used to analyze the overall survival (OS) probabilities between high-risk and low-risk groups using R Survival^21^ and R survivalROC^22^, respectively. The test dataset and an additional external dataset for blind validation were used to evaluate the prognostic value of the developed model.

### Nomogram construction

Since miRNA-based risk score was unlikely to be the only prognostic predictor for overall survival, the clinical profile was also considered. Both univariate and multivariate Cox regression analyses were performed with some clinical features, namely age, pregnancies, smoking_history, grade, stage, and HPV_status. Only those clinical variables that survived both the analyses were used with the miRNA-based risk score to build an integrated nomogram map that tabulates the probability of one-year and five-year OS of CESC. The discrimination was quantified using Harrell’s concordance index (C-index), and calibration performed using bootstrap with 1000 resamples.

## RESULTS

The TCGA expression data consisted of expression values of 2589 miRNA in 312 samples enrolled in this study, including 309 cervical cancer tissues and 3 matched normal tissues. Post data preprocessing, we obtained an expression dataset consisting of 467 miRNAs across 303 samples with stage annotation. Table 1 shows the distribution of samples according to the AJCC staging system^23^. The demographic features and clinical characteristics considered, namely age, smoking history, vital status, pregnancies, HPV status, and histologic grade are summarized in Table 2. Fitting the linear model and applying the filter criteria yielded a total of 101 differentially expressed miRNAs between cervical cancer tissues and matched normal tissues, provided in Supplementary File S1. Most of the top-ranked miRNAs are overexpressed (for e.g, hsa-miR-200c-3p, hsa-miR-141, hsa-miR-200a, hsa-miR-21-5p), suggesting oncomir function with increased epigenetic suppression of target tumor-suppressor gene expression. Table 3 shows the top ten miRNAs with their stage-wise log2FC and linear model significance.

**Table 1.**
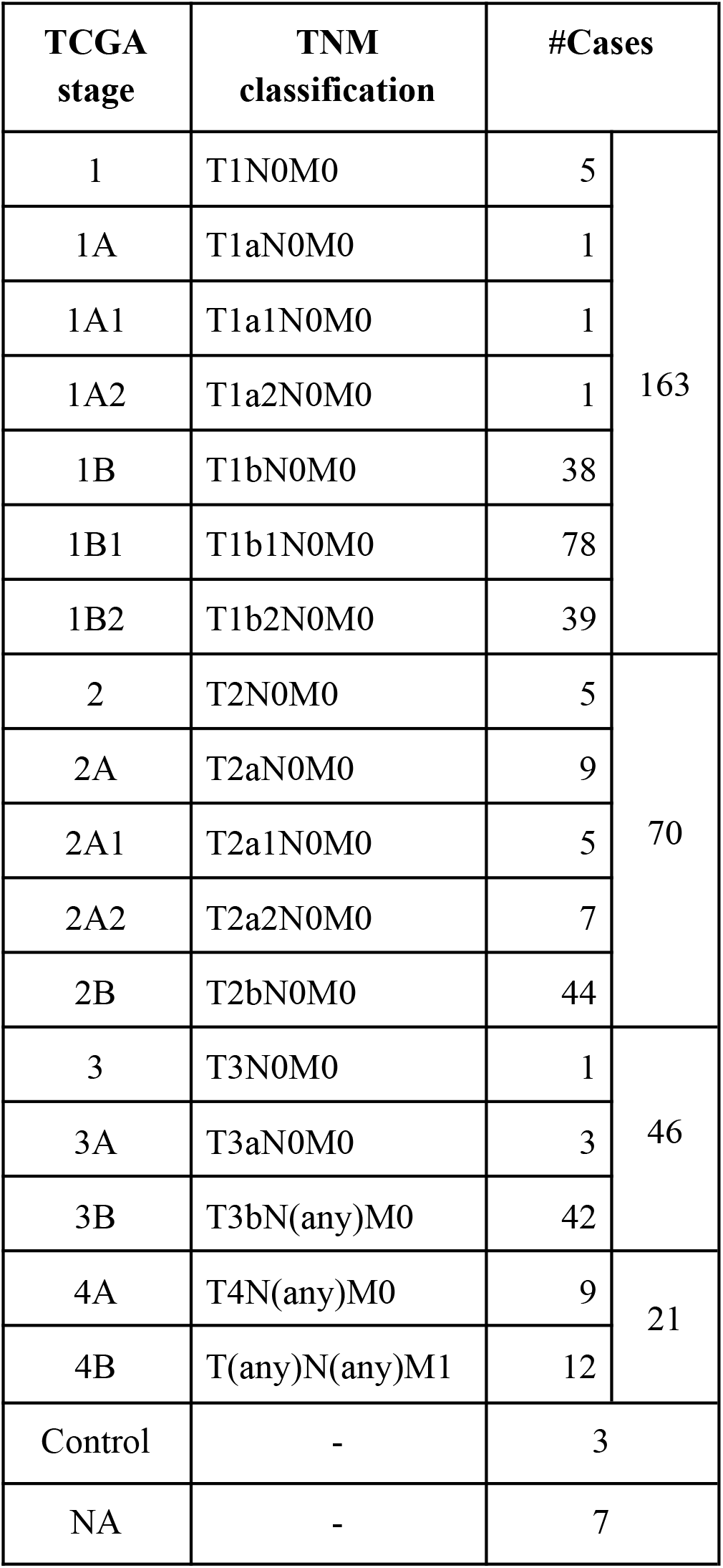
Distribution of cases by stage. AJCC staging is represented by the TNM (Tumor-Node-Metastasis) code. Control refers to matched normal samples, and ‘NA’ denotes cases with unavailable stage information.

| TCGA stage | TNM classification | #Cases |  |
| --- | --- | --- | --- |
| 1 | T1N0M0 | 5 | 163 |
| 1A | T1aN0M0 | 1 |  |
| 1A1 | T1a1N0M0 | 1 |  |
| 1A2 | T1a2N0M0 | 1 |  |
| 1B | T1bN0M0 | 38 |  |
| 1B1 | T1b1N0M0 | 78 |  |
| 1B2 | T1b2N0M0 | 39 |  |
| 2 | T2N0M0 | 5 | 70 |
| 2A | T2aN0M0 | 9 |  |
| 2A1 | T2a1N0M0 | 5 |  |
| 2A2 | T2a2N0M0 | 7 |  |
| 2B | T2bN0M0 | 44 |  |
| 3 | T3N0M0 | 1 | 46 |
| 3A | T3aN0M0 | 3 |  |
| 3B | T3bN(any)M0 | 42 |  |
| 4A | T4N(any)M0 | 9 | 21 |
| 4B | T(any)N(any)M1 | 12 |  |
| Control | - | 3 |  |
| NA | - | 7 |  |

**Table 2.** Clinical profile of cervical cancer patients. Summary of key clinical / demographic features of the dataset. For ordinal / continuous variables (age, smoking_history, and pregnancies), the mean ± standard deviation is given. Histologic grade refers to the degree of differentiation in the cancer sample. It is seen that most cervical cancer patients present with HPV+ status.

| Characteristic |  | StageI | StageII | StageIII | StageIV | 'NA' | Overall |
| --- | --- | --- | --- | --- | --- | --- | --- |
| Number of samples |  | 163 | 70 | 46 | 21 | 7 | 307 |
| Age (years) | | 45.9±13.2 | 49.1±14.2 | 51.2±13.4 | 53.3±12.6 | 58.8±18.8 | 48.3±13.8 |
| HPV status | Positive | 152 | 63 | 44 | 18 | 7 | 284 |
|  | Negative | 11 | 6 | 2 | 3 | - | 22 |
|  | Indeterminate | - | 1 | - | - | - | 1 |
| Smoking history | | 1.8±1.1 | 1.7±1.2 | 1.9±1.1 | 1.7±1.1 | 2.7±2.1 | 1.8±1.2 |
| Pregnancies | | 3.3±2.1 | 3.9±3.1 | 4.1±2.8 | 3.7±2.4 | 2.5±2.1 | 3.6±2.6 |
| Vital status | Alive | 135 | 61 | 36 | 8 | 7 | 247 |
|  | Dead | 28 | 9 | 10 | 13 | - | 60 |
| Histologic Grade | G I/II | 84 | 34 | 23 | 11 | 2 | 154 |
|  | G III/IV | 65 | 27 | 20 | 5 | 4 | 121 |

**Table 3.**
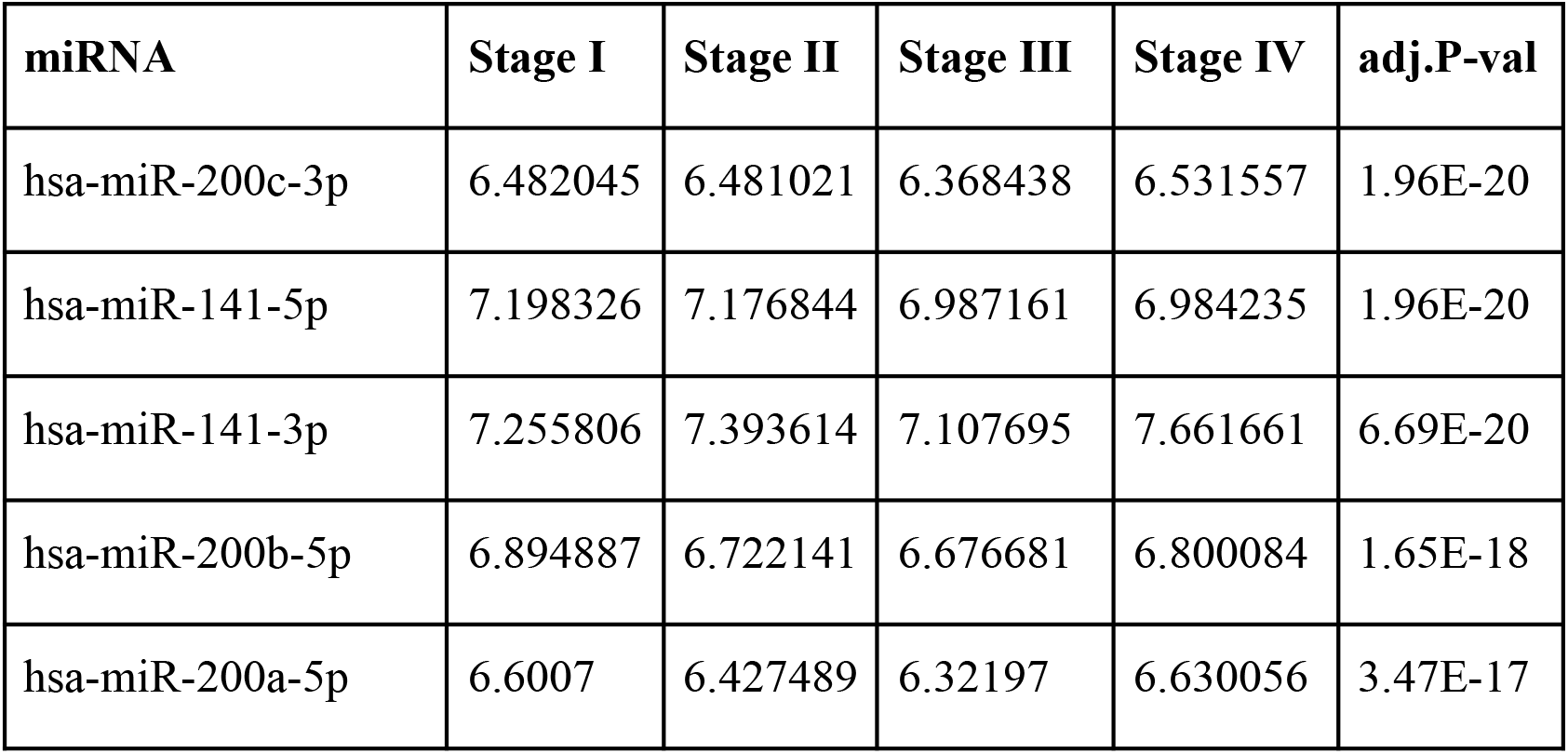
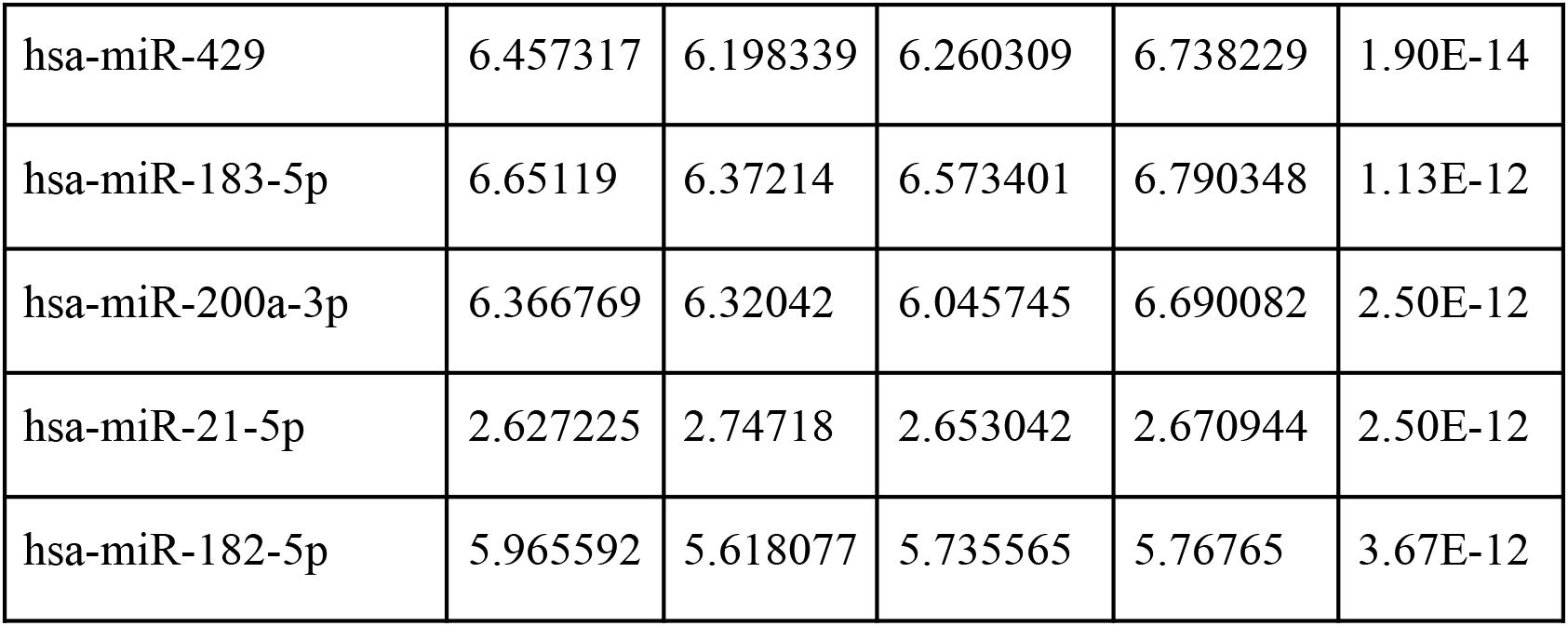
Top 10 miRNAs of the linear model. The log-fold change expression of the miRNA in each stage relative to the controls is given, followed by p-value adjusted for multiple hypothesis testing.

Each DEmiR was subjected to univariate Cox modeling to evaluate its prognostic significance. This process identified only 52 miRNAs as significantly associated with overall survival, based on p-value < 0.05 (data presented in Supplementary File S2). To optimize the dimensions of the prognostic miRNA biomarker panel, we applied Lasso-penalized Cox regression on the 52 miRNAs to obtain five miRNAs, hsa-miR-625-5p, hsa-miR-3934-5p, hsa-miR-330-3p, hsa-miR-642a-5p, has-miR-95-3p, Only one miRNA, hsa-miR-616-5p, survived the SVM-RFE feature selection process. Figure 2 shows the union of these results (i.e, the six miRNAs).

**Figure 2.**
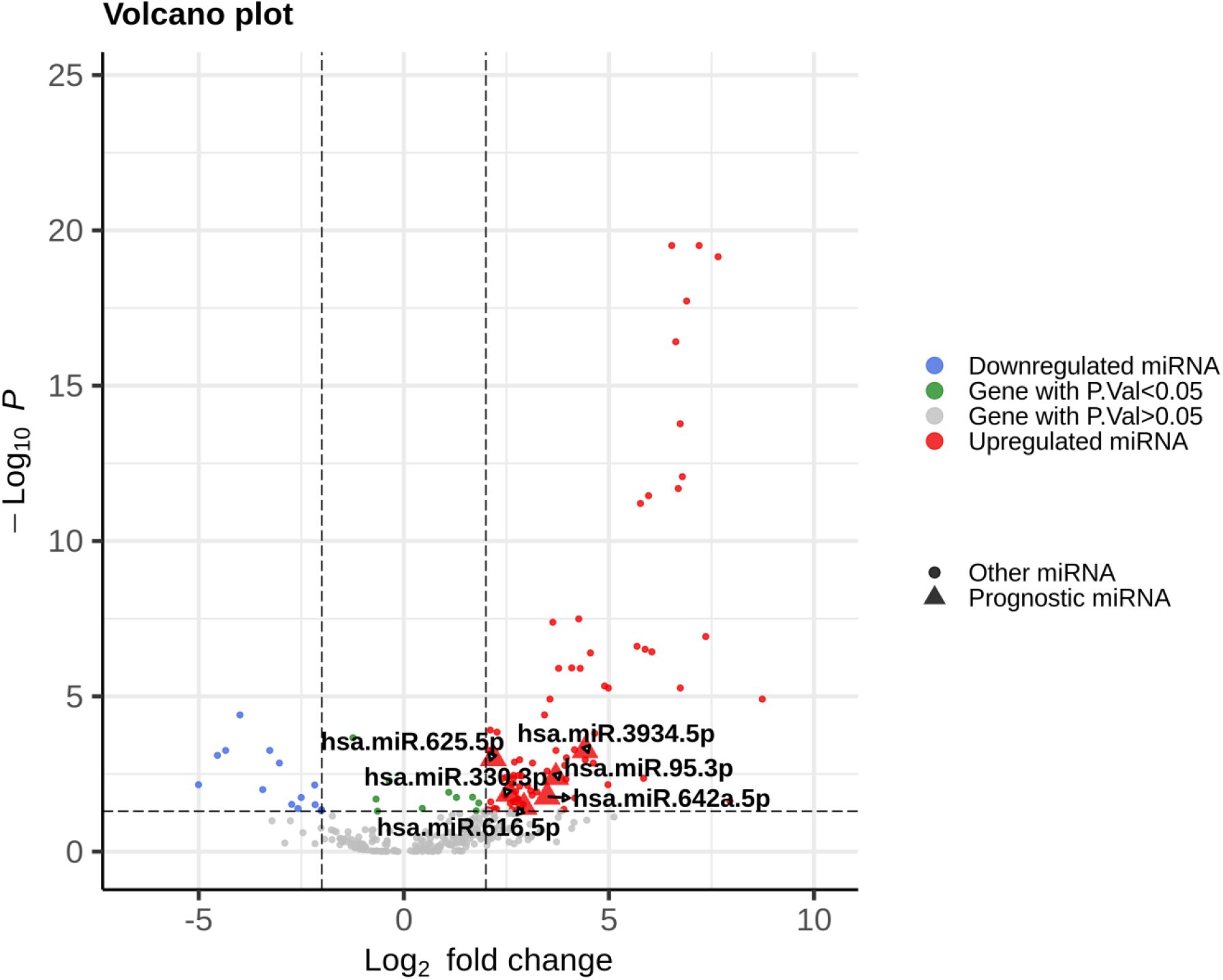
Volcano plot of the expression distribution of the miRNAs with non-trivial expression in the dataset, highlighting the upregulated and downregulated DEmiRs, and the prognostic DEmiRs post the feature selection process. Interestingly, all the prognostic DEmiRS were upregulated, but none were an outlier DEmiR (top right). X-axis denotes log2(FC) of expression with respect to control, and the Y-axis denotes the -log10 transformation of the p-value significance of the linear model for the respective miRNA.

The six miRNAs were taken forward for multivariate survival analysis, and subjected to a stepwise backward-selection process, to further compact the miRNA signature. This process yielded an optimal signature of three miRNAs namely hsa-miR-625-5p, hsa-miR-330-3p, and hsa-miR-95-3p, with model p-value < 0.002 (Table 4), for construction of the CESC prognostic risk model.

**Table 4.** Summary of the results of CESC Cox analysis. It is seen that hsa-miR-625-5p has a significant protective effect on CESC OS, in contrast with hsa-miR-95-3p and hsa-miR-330-3p. The overall multivariate model is very significant with p-value < 0.002. HR denotes hazard rate, and CI confidence interval.

| Variables | Analysis | Coefficient | HR (95% CI) | P-value |
| --- | --- | --- | --- | --- |
| hsa-miR-95-3p | Univariate | -0.84 | 0.43 (0.24-0.79) | 0.0063 |
|  | Multivariate | 0.30 | 1.35 (1.05-1.73 ) | 0.0197 |
| hsa-miR-625-5p | Univariate | 1.4 | 4.2 (1.3-14) | 0.0180 |
|  | Multivariate | -0.52 | 0.59 (0.43-0.83) | 0.0020 |
| hsa-mir-330-3p | Univariate | -0.68 | 0.51 (0.28-0.93) | 0.0290 |
|  | Multivariate | 0.35 | 1.42 (0.98-2.03) | 0.0608 |

The CESC prognostic risk model, given by eqn. (1), was then parameterized using the expression of these three miRNAs:

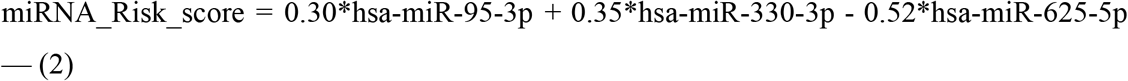

It is seen that hsa-miR-625-5p has a significant protective effect on CESC OS, whereas the expression of hsa-miR-95-3p and hsa-miR-330-3p elevate the risk. Based on this model, we computed the risk score for each patient in the train dataset, and used the maxstat of the resulting risk-score distribution to separate patients into high- and low-risk groups (Figure 3A). The Kaplan–Meier survival curve of this distribution revealed significantly worse prognosis in the high-risk group (p-value < 1E-4) (Figure 3B). Time-dependent ROC analysis of the risk-score model on the train dataset for 1-, 2-, 3-, and 5-year overall survival yielded prognostic AUC values of 0.71, 0.72, 0.74 and 0.73, respectively (Figure 3C). These results encouraged validation of the CESC-related prognostic signature on the test dataset, whose risk-score distribution is shown in Figure 4A. The following outcomes validated the results: (i) Kaplan-Meier survival curve showed significantly worse prognosis in the high-risk group (p-value < 1E-4) (Figure 4B) ; and (ii) time-dependent AUROC values 0.84, 0.79, 0.71 and 0.71 were obtained for 1-, 2-, 3-, and 5-year overall survival, respectively (Figure 4C).

**Figure 3.**
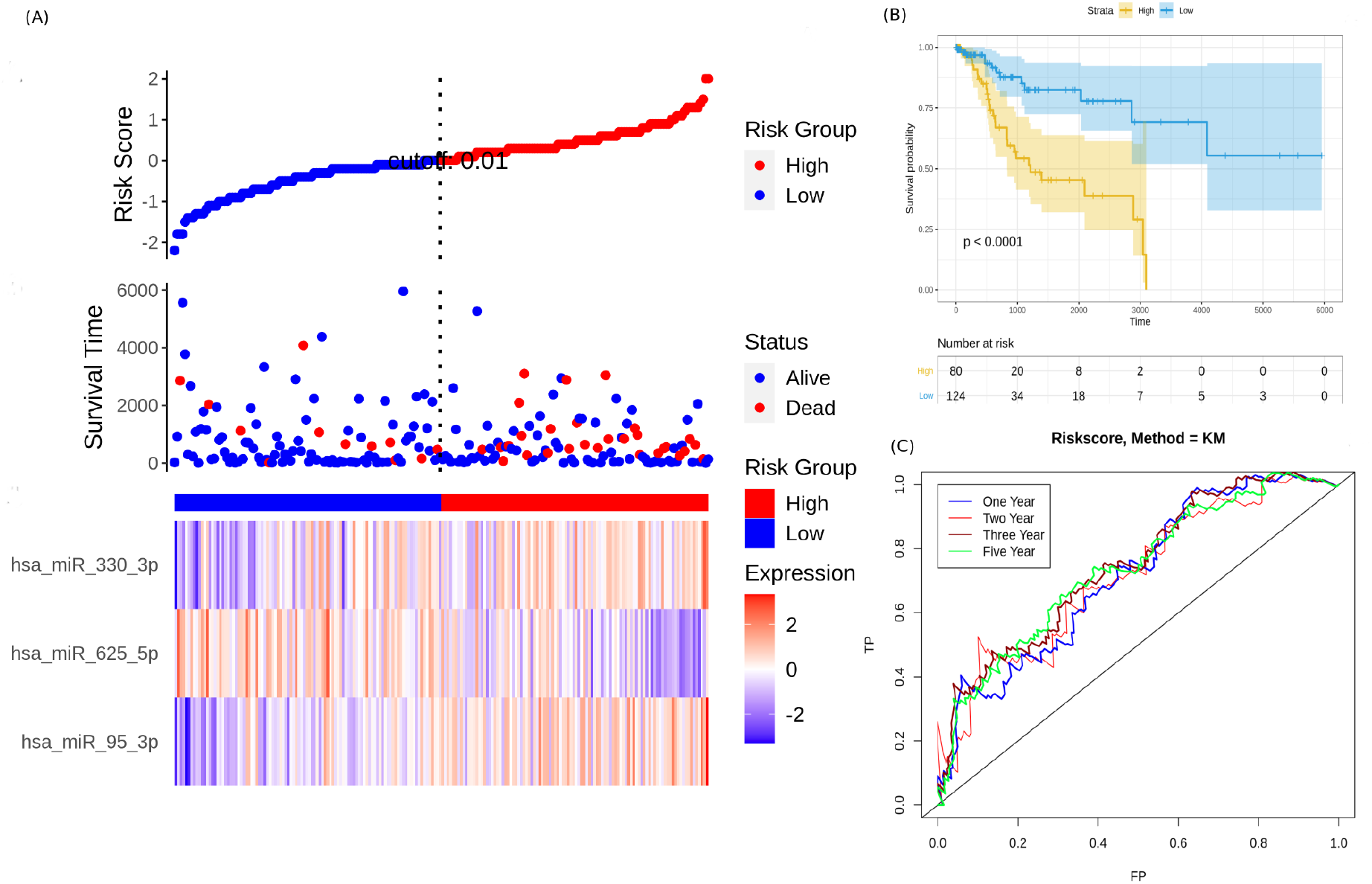
Performance of the constructed risk-score model on train dataset. (A) This panel shows the risk-score value (top), survival status (middle), and expression of the three prognostic miRNAs (bottom) for each patient, sorted by the risk-score distribution. Patients were stratified into low-risk (blue) and high-risk (red) groups according to the risk-score value. The patterns in the expression profiles accord with the signed risk of the respective miRNAs. (B) Kaplan–Meier survival curves based on the three-miRNA prognostic signature showing significant difference between the two groups. (C) Time-dependent ROC curves for 1-, 2-, 3-, and 5-year overall survival predictions using the given model.

**Figure 4.**
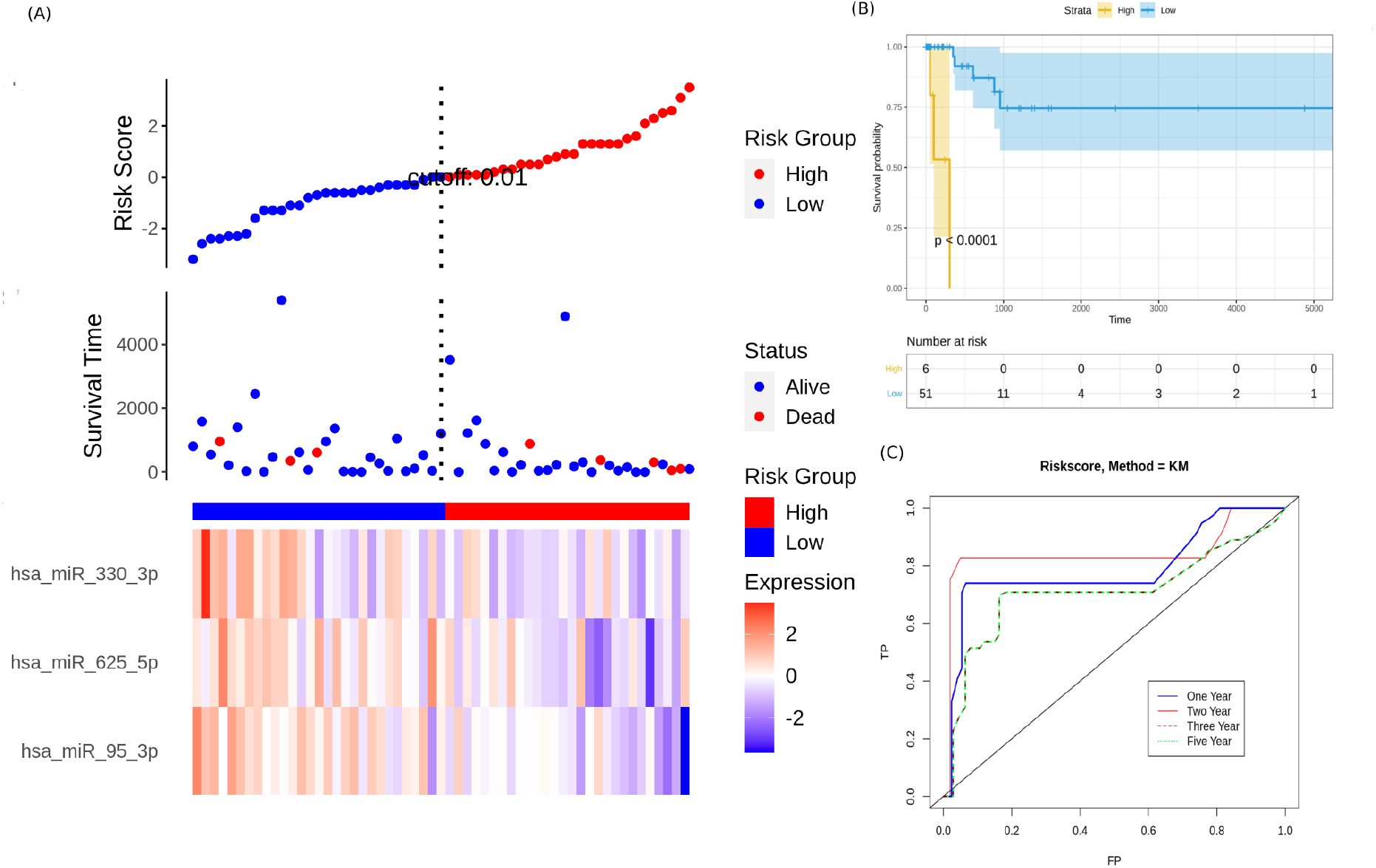
Performance evaluation of the constructed risk-score model on unseen test dataset. (A) This panel shows the risk-score value (top), survival status (middle), and expression of the three prognostic miRNAs (bottom) for each patient, sorted by the risk-score distribution. Patients were stratified into low-risk (blue) and high-risk (red) groups according to the median risk-score value. (B) Kaplan–Meier survival curves based on the three-miRNA prognostic signature showing significant difference between the two groups. (C) Time-dependent ROC curves for 1-, 2-, 3-, and 5-year overall survival predictions using the given model.

To validate the prognostic value of the model on an external, out-of-domain dataset, we used the study results of How et al^24^. This study used a TaqMan Low Density Array (TLDA) to measure expression in formalin-fixed paraffin-embedded (FFPE) cervix samples. Two datasets from the study were used:

1. Normalized and log_2_-transformed miRNA expression data of 87 FFPE cervix samples used for validation, available at: https://www.ncbi.nlm.nih.gov/pmc/articles/PMC4399941/bin/pone.0123946.s005.txt; and
2. corresponding clinical information, available at: https://www.ncbi.nlm.nih.gov/pmc/articles/PMC4399941/bin/pone.0123946.s002.xlsx. The clinical information was used to annotate the samples, and the expression subset corresponding to the three miRNAs in the optimal risk model (i.e. eqn. 2) was extracted. Since the miRNA arm information (−3p or -5p) was missing for hsa-miR-625 and hsa-miR-95, the arm-neutral expression values for both these miRNAs were used. The risk score for each sample was calculated based on eqn. 2, and the resulting risk score distribution was stratified into high-risk and low-risk patient groups based on the maxstat statistic computed by R survminer. The curves were visualized using Kaplan-Meier analysis, yielding significantly worse prognosis (*P* < 0.032) in the high-risk patient group relative to the low-risk group (Figure 5).

**Figure 5.**
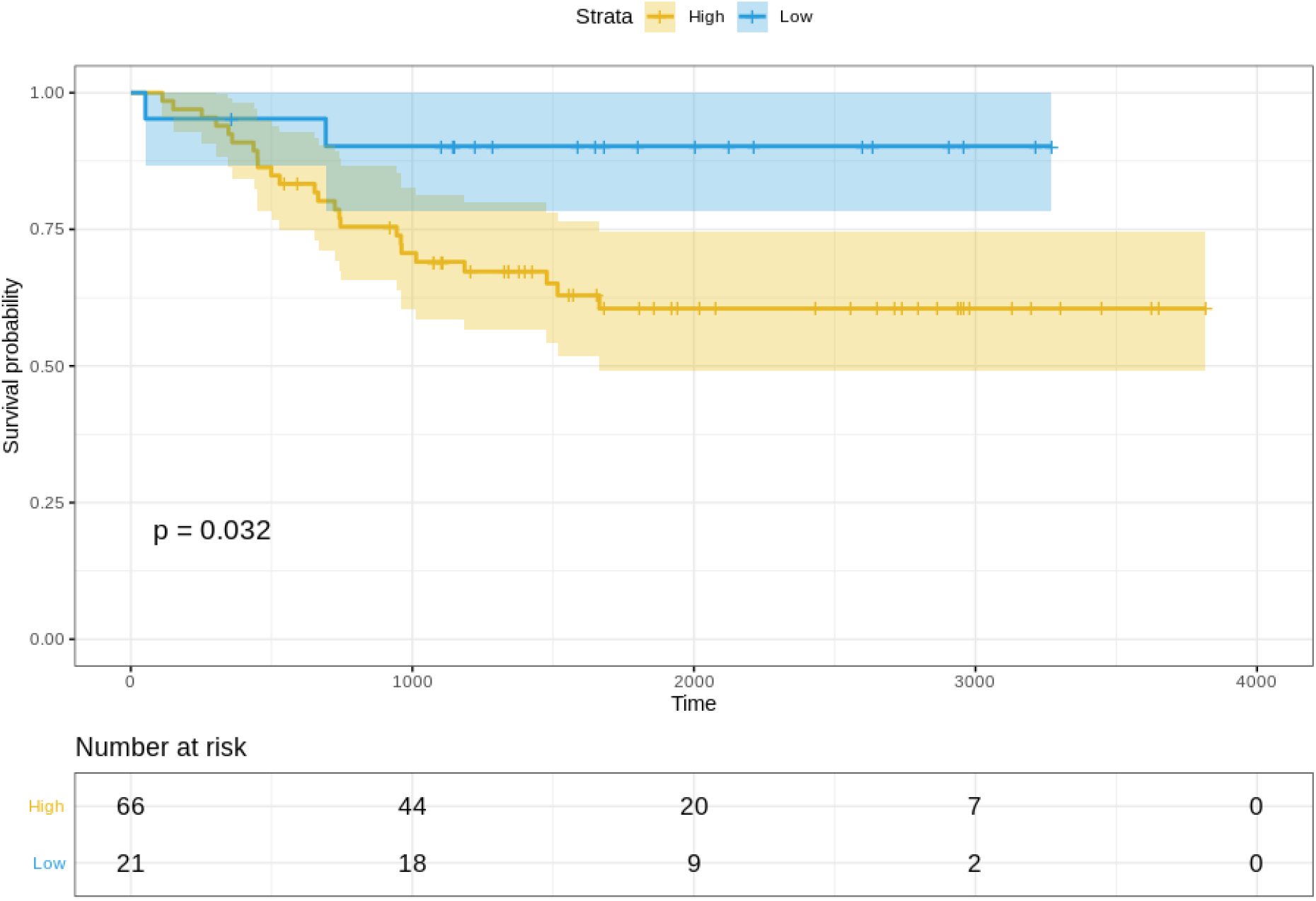
Kaplan–Meier survival curves for the validation dataset, showing significantly worse prognosis for the high-risk patient group relative to the low-risk group. 95% confidence bands for the risk groups are also shown.

Certain clinical features namely age, HPV_status, pregnancies, smoking_history, histologic_grade, and stage could boost the prognostic predictive value, and hence were examined for candidate inclusion in the risk model. Each clinical feature was subjected to the univariate Cox survival analysis, and only one clinical feature turned out significant, namely the stage. This was used with the miRNA-based risk-score to model an integrated multivariate Cox logistic regression. Both the factor levels of both the variables were significant, and the overall multivariate model was extremely significant (p-value ∼ 4E-05) (Figure 6). The integrated CESC prognostic risk model was then parameterized as:

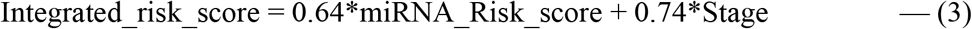

**Figure 6.**
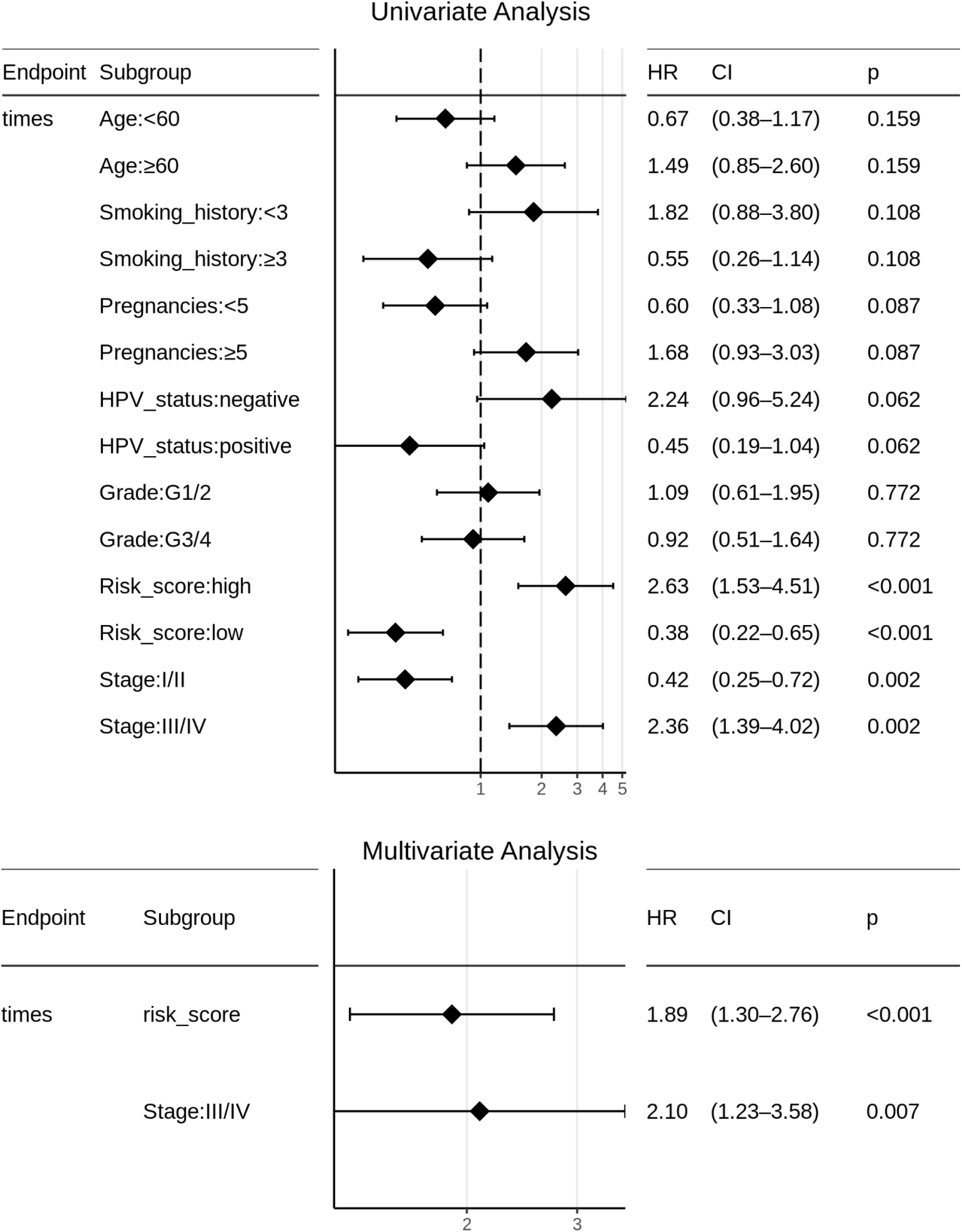
Univariate and multivariate Cox logistic regression analyses of patient clinical profile, with respect to CESC OS. Surprisingly, patient HPV-status is not significant to CESC OS, and the tumor Grade is almost irrelevant to prognosis here. Note that both the levels of clinical stage (viz. Stage:I/II and Stage:III/IV) are significant, and constitute an independent risk factor in addition to miRNA_risk_score.

Based on the risk models developed, a nomogram was built to predict one-year and five-year survival probabilities (Figure 7). The nomogram C-index was estimated as 0.7136 ± 0.047, indicating good discrimination. Further, the nomogram calibration plots for one-year and five-year OS probabilities based on bootstrap resampling showed consistency between the predicted and actual survival probabilities (Figure 8).

**Figure 7.**
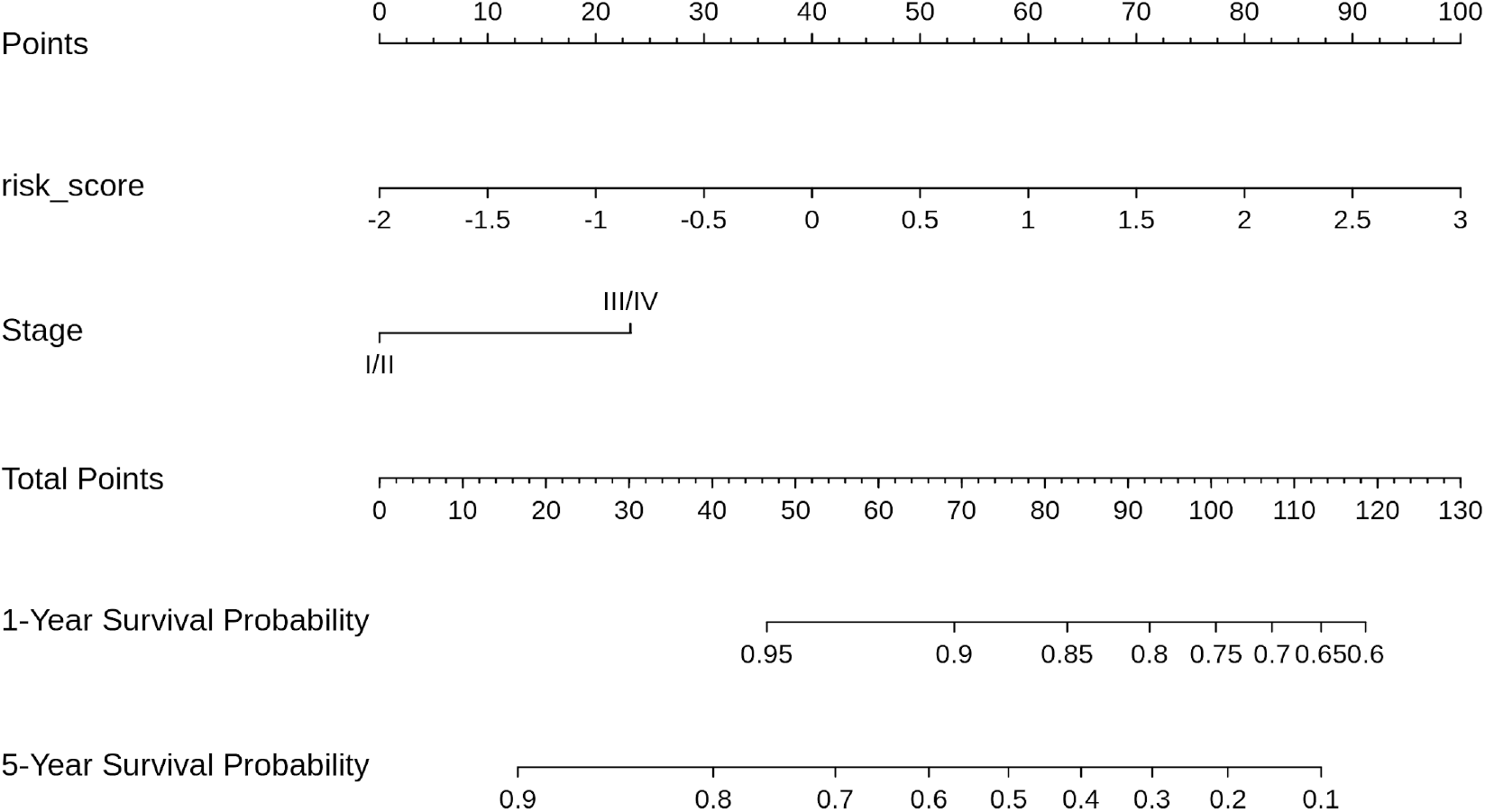
Nomogram for reading the overall survival in CESC sample, according to miRNA_risk_score (eqn. 2) and clinical stage.

**Figure 8.**
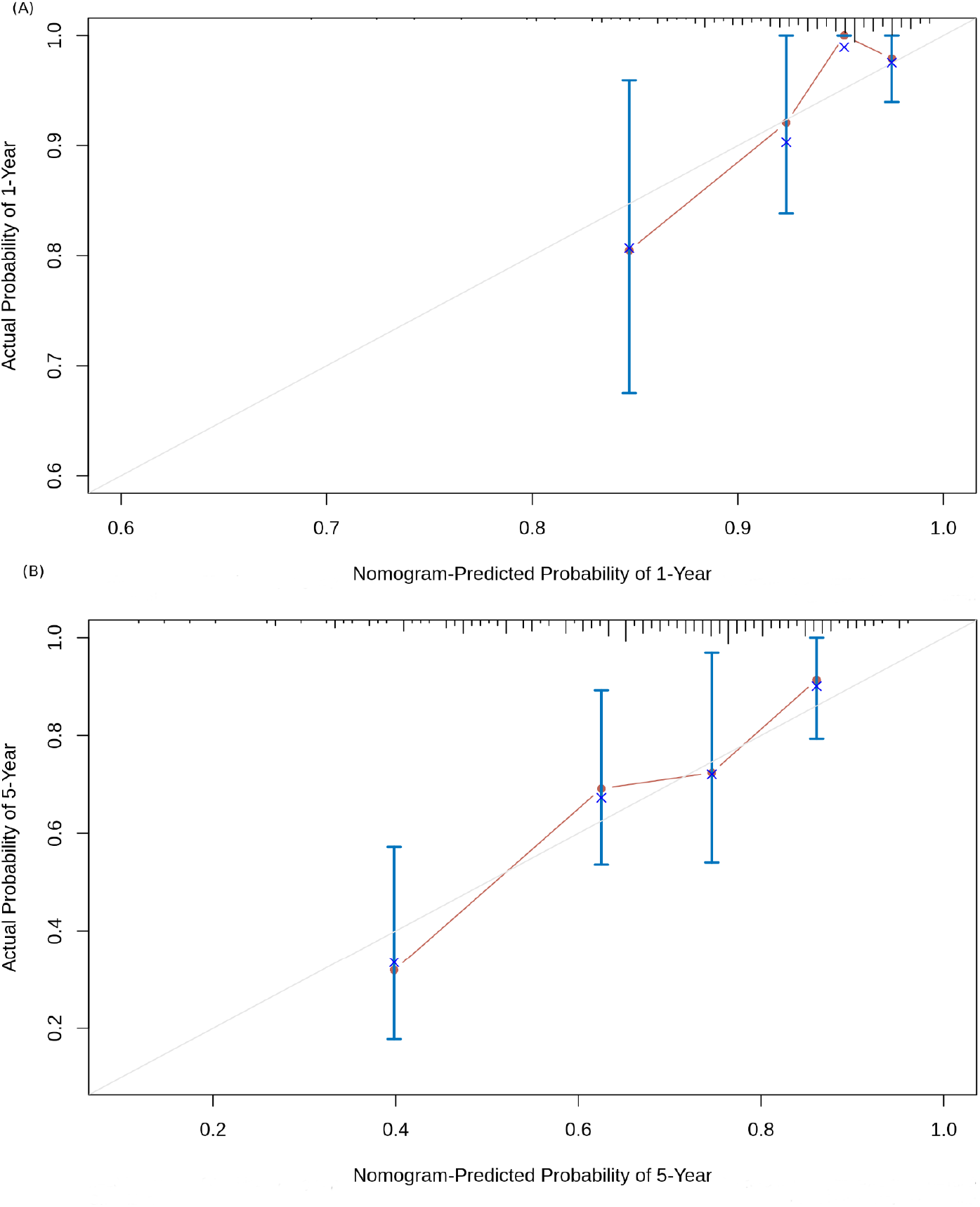
Nomogram calibration curves. (A) 1-year OS probability; (B) 5-year OS probability. The four sub-cohorts of the dataset are visualized, and the corresponding x represents the bootstrap-corrected estimates of the nomogram performance along with the standard error. The solid line compares the nomogram performance with the reference truth.

## DISCUSSION

MiRNAs add a layer of critical regulatory control over genomic expression, and aberrations in their expression could lead to the development of cancer hallmarks^25^. MiRNAs could be detected in the serum, and lend valuable potential as diagnostic and prognostic biomarkers of various cancers, including cervical cancer^26^. Several prognostic miRNAs for CESC have been reported, including miR-31^27^, miR-155^28^, and miR425-5p^29^. However a systematic hypothesis-free scan for comprehensive miRNA signatures remains missing in the literature. In this study, we have attempted to fill this void with an integrated multi-layered bioinformatics approach to the detection of a reliable prognostic DEmiR biomarker signature. The study has yielded three prognostic miRNAs, namely hsa-miR-625-5p, hsa-miR-95-3p, and hsa-miR-330-3p. Downregulation of hsa-miR-625-5p has been documented in many cancers including bladder cancer^30^, non-small cell lung cancer^31^, hepatocellular carcinoma^32^, melanoma^33^ and cervical cancer^34^. A causal mechanism relating miR-625-5p expression to inhibition of cervical cancer cell growth via suppression of NF-κB signaling has been reported^35^, consistent with its mirsupp identity disclosed here. Sponging miR-625-5p in turn is likely to drive cervical cancer progression, and this has been demonstrated recently^36^. Jafarzadeh et al. suggested that miR-330-3p promoted pro-tumorigenic events in various cancers like lung cancer, pancreatic cancer, bladder cancer and cervical cancer, and that its downregulation could stall tumor development^37^, both observations consistent with its oncomir identity disclosed here. Further, miR-95-3p has been implicated in activating the wnt/βcatenin pathway in prostate cancer tissues^38^, thereby promoting cell proliferation, migration and invasion, consistent with its oncomir identity disclosed here.

To examine the network-level effects of these miRNAs, we retrieved the RNA-Seq transcriptome for each patient in our dataset from firebrowse.org, and correlated this data with the expression of the three miRNAs of interest to infer potential target genes. Target genes with substantial inverse correlation in expression (defined as Pearson ρ or Spearman ρ or Kendall τ < -0.3) were identified, and the consensus with multiMiR^39^ predictions for each of the three miRNAs was investigated. This yielded three consensus target genes with respect to hsa-miR-95-3p, namely NXPH3, BOC, EID1; two consensus target genes with respect to hsa-miR-625-5p, namely SIN3B and TPRG1L; and two consensus target genes with respect to hsa-miR-330-3p, namely THRA and DYRK2. Functional enrichment analysis of the consensus genes conducted with miRNeT^40^ on GO and KEGG databases yielded significance for cancer pathways and cell cycle regulation. We also used the miR2Trait server^41^ to investigate the diseasome of this three-miRNA signature, and found significance for ‘uterine cervical neoplasm’ (p-value ∼1.5E-3), ‘squamous cell carcinoma’ (p-value ∼7.7E-3), and ‘cervical intraepithelial neoplasia’ (p-value ∼2.2E-2). Detailed results of the above investigations are presented in Supplementary File S3.

Nomograms are widely used for simplifying the task of interpretation from models, and have been constructed with miRNAs for cervical cancer screening^42^, prognosis^43^, and recurrence risk^44^. To facilitate the ready prognosis of cervical cancer patients, the models developed in this work were re-built with the full (train + test) dataset, and served as a web-app named CESCProg, deployed at: https://apalania.shinyapps.io/cescprog/ for non-commercial uses. The concerned user may provide the form inputs, namely the expression values of the three prognostic DEmiRs and an optional sample staging information. Based on the user request, the app proceeds to classify the risk of the sample, and compute a risk-score based on eqn. 2 or eqn. 3. The calculated risk-score is then consulted with the back-end nomogram to estimate the one-year and five-year survival probabilities. Serum-based or cervical mucus-based miRNAs are minimally invasive, and could be detected and quantified using a range of techniques (for e.g, see ref. 45).

## CONCLUSIONS

MiRNA biomarkers are an emerging diagnostic and prognostic aid to the management of disease, especially cancers. Here we present CESCProg, an miRNA-based prognostic model for cervical cancer developed by applying a sequence of purifying filters to the TCGA CESC dataset. All the three miRNAs in the panel, namely hsa-miR-95-3p, hsa-miR-330-3p and hsa-miR-625-5p, show upregulation in cervical cancer relative to controls, suggesting feasibility for detection as biomarkers. In the miRNA risk model, hsa-miR-625-5p exhibits a protective effect on OS, while the other two miRNAs elevate the risk. The miRNA risk model was effective and extremely significant in stratifying CESC OS on the test dataset. A second risk model was developed with the inclusion of clinical features to maximize nomogram discrimination. This yielded a C-index of 0.7136 ± 0.047. The models have been deployed as a web-service as a possible aid to medical decision-making. They are available for non-profit use at: https://apalania.shinyapps.io/cescprog.

## Data Availability

https://doi.org/10.6084/m9.figshare.21543150.v2

## ACKNOWLEDGMENTS

We would like to thank the School of Chemical and Biotechnology & CeNTAB, SASTRA Deemed University, for computing and infrastructure support.

